# Multi-omics liquid biopsy identifies mitochondrial dysfunction in geographic atrophy and supports the longevity-associated metabolite α-ketoglutarate as a therapeutic strategy

**DOI:** 10.64898/2026.03.12.26347263

**Authors:** Tsai-Chu Yeh, Gabriel Velez, Architesh Prasad, Soo Hyeon Lee, Ditte K. Rasmussen, Aarushi Kumar, Madhumeeta Chadha, Mohamed Ziad Dabaja, Aneal M Singh, Steven Sanislo, Stephen Smith, Prithvi Mryuthyunjaya, Artis Montague, Alexander G. Bassuk, David Almeida, Antoine Dufour, Vinit B Mahajan

## Abstract

**Background:** Mitochondrial dysfunction is an emerging metabolic hallmark of age-related diseases, yet tools to directly profile mitochondrial pathways and test metabolic interventions in the living human eye remain limited. Multi-omics ocular liquid biopsy enables real-time proteomic and metabolomic profiling of the intraocular microenvironment, complementing systemic biomarkers and imaging surrogates. Here, we used this approach to define mitochondrial and tricarboxylic acid (TCA) cycle dysregulation in geographic atrophy (GA) and to assess whether oral α-ketoglutarate (α-KG) supplementation can modulate mitochondrial metabolites within the eye.

**Methods:** Mitochondrial and TCA cycle-related proteins were profiled in aqueous humor (AH) samples from patients with GA using DNA-aptamer-based proteomics. In a phase 0 study, a second cohort undergoing sequential cataract surgery provided paired AH samples collected at first-eye surgery and at second-eye surgery after interim α-KG supplementation. These samples underwent targeted metabolomic profiling using hydrophilic interaction liquid chromatography coupled with mass spectrometry.

**Results:** In GA, 64 mitochondrial proteins were differentially expressed, including coordinated TCA-cycle deficiencies marked by reduced expression of enzymes regulating TCA entry and flux, including PDHB and DLST. In the phase 0 cohort, oral α-KG supplementation significantly increased intraocular α-KG levels and the α-KG-to-succinate ratio (P < 0.05), with coordinated shifts across TCA intermediates consistent with enhanced TCA cycle flux.

**Conclusions:** AH proteomics demonstrated mitochondrial pathway depletion in GA, consistent with reduced oxidative bioenergetic capacity. AH metabolomics provided first-in-human in vivo evidence that systemic α-KG supplementation can modify intraocular metabolites and may enhance intraocular energy metabolism. These findings support ocular liquid biopsy as a precision-health framework for per-patient biomarker-guided metabolic trials in GA.

**Plain Language Summary:** Geographic atrophy (GA) is an advanced form of age-related macular degeneration and a major cause of irreversible vision loss. To better understand the biology of GA, we studied proteins and small molecules in aqueous humor, the fluid inside the eye. We found that eyes with GA showed clear signs of mitochondrial dysfunction, including disruptions in the tricarboxylic acid (TCA) cycle, a key pathway for energy production. This suggests that impaired cellular metabolism is an important feature of the disease. We then tested whether oral α-ketoglutarate (α-KG), a metabolite involved in mitochondrial function and previously shown to have life-extending effects in preclinical studies, could alter these metabolic pathways in the human eye. We found that α-KG supplementation not only increased intraocular α-KG levels but changed metabolic markers linked to mitochondrial activity, providing the first direct evidence that oral supplementation can reach the eye and measurably modify metabolism inside the living human eye. Together, these findings show that liquid biopsy can provide a direct molecular snapshot of the living human eye and may help accelerate the development of biomarker-guided therapies for ocular diseases.

**Key Points:** *Questions:* What specific mitochondrial and TCA-cycle dysfunctions occur in the aqueous humor (AH) of patients with geographic atrophy (GA), and can oral α-ketoglutarate (α-KG) supplementation measurably remodel these metabolic pathways in the living human eye?

*Findings:* AH proteomics in GA patients revealed significant mitochondrial disruption and a coordinated depletion of TCA-cycle enzymes. In a paired-eye interventional metabolomics study, oral α-KG significantly increased intraocular α-KG levels and the α-KG-to-succinate ratio, proving that systemic therapy can drive measurable metabolic modulation within the human eye.

*Meaning:* Multi-omics liquid biopsy provides a direct, eye-specific readout of mitochondrial metabolism in GA and offers early human proof-of-concept that a systemic metabolic therapy can successfully reach and modify intraocular pathways, paving the way for biomarker-guided clinical trials in AMD.

## Introduction

Geographic atrophy (GA), the advanced form of non-exudative age-related macular degeneration (AMD), is a major cause of irreversible vision loss for which current anti-complement therapies only show modest effects on structural progression.^1,2^ This highlights the need to define alternative pathogenic mechanisms and molecular pathways that may contribute to disease progression and inform therapeutic opportunities. Across diverse eye diseases, including AMD, emerging proteomic and metabolomic evidence points to a shared mitochondrial dysfunction, including reduced tricarboxylic acid (TCA) cycle activity, impaired oxidative phosphorylation, and altered redox regulation.^3–6^ Within AMD specifically, broader metabolic dysregulation, most notably abnormalities in lipid and amino acid metabolism, has also been implicated .^7–10^ However, much of this evidence is derived from systemic (plasma-based) metabolomic studies, and whether GA exhibits similar mitochondrial pathway dysregulation directly within the human eye remains unresolved. This knowledge gap is also especially important given the long-standing emphasis on oral antioxidant nutritional supplements for AMD, including the AREDS and AREDS2 formulations.^11,12^ Although these trials demonstrated clinical benefit in earlier AMD stages, they were not designed to measure intraocular metabolic effects.

Prior human aqueous humor (AH) studies in age-related ocular diseases have reported mitochondrial pathway perturbations, including reduced α-ketoglutarate (α-KG) levels, suggesting a disease-associated metabolic deficit within the eye.^13,14^ α-KG is a key intermediate of the TCA cycle and participates in multiple interconnected metabolic pathways. By providing anaplerotic input to sustain mitochondrial TCA flux and energy production, α-KG supports a core bioenergetic function. In addition, α-KG contributes to cellular redox balance and modulates oxygen-dependent metabolic programs, positioning it as a central regulator of mitochondrial homeostasis and stress-response pathways.^15,16^ In preclinical models, previous studies show that augmenting α-KG availability enhances mitochondrial resilience, attenuates inflammaging, and extends lifespan, and our prior work demonstrates that α-KG confers neuroprotective effects in models of retinal degeneration.^17–19^ Together, these findings provide a mechanistic rationale for targeting α-KG specifically, rather than other TCA intermediates.

Liquid-biopsy proteomics and metabolomics allows direct molecular interrogation of the living human eye. To date it remains unexplored whether such an approach can measure mitochondrial proteins and their metabolites in vivo, enabling molecular pathway-level analyses that were previously restricted to postmortem or animal studies.^20^ If feasible, multi-omics liquid biopsies could serve as a precision health platform for systematically investigating metabolic therapeutic interventions. In this study, we performed AH proteomic profiling to determine whether mitochondrial pathways, including the TCA cycle, are disrupted in GA patients. We also tested whether oral α-KG supplementation can, through mass-action effects, partially offset mitochondrial enzymatic constraints.

## Methods

### Study design and participants

This single-center, prospective study consisted of two complementary participant groups: (1) patients with GA undergoing AH collection for proteomic profiling, and (2) patients undergoing sequential bilateral cataract surgery for a metabolic intervention with oral calcium α-ketoglutarate (Ca-AKG). All study procedures were approved by the Stanford University Institutional Review Board, complied with HIPAA regulations, adhered to the Declaration of Helsinki, and written informed consent was obtained from all participants. This study was registered at ClinicalTrials.gov (NCT07269704).

### AH sample collection in GA

Twenty patients with clinically confirmed non-exudative late AMD with well-defined GA were recruited during outpatient ophthalmic evaluations for proteomics analysis. Exclusion criteria included prior intraocular surgery other than uncomplicated cataract extraction, active ocular inflammation, or coexisting retinal disease. Approximately 50 μL of AH was aspirated from the anterior chamber using a sterile 30-gauge needle. Samples were placed immediately on ice, aliquoted, and stored at -80 °C until proteomic analysis.

### Aptamer-based proteomics analysis

For proteomic profiling, AH samples were shipped on dry ice overnight to Somalogic (Boulder, Colorado, USA) and the samples were analyzed with an aptamer-based proteomics assay (SomaScanⓇ Assay v4.1, Somalogic) as previously described. Briefly, expression levels of 695 human mitochondrial and TCA protein targets selected by gene ontology were evaluated by corresponding DNA aptamers, which are short single-stranded, chemically modified DNA molecules which specifically bind to their protein targets and which are quantified using DNA microarray technology.

### AH sample collection and Ca-AKG supplementation in sequential bilateral cataract surgery patients

A second cohort of eight participants scheduled for sequential bilateral phacoemulsification with intraocular lens implantation were enrolled. Inclusion criteria were age >50 years, planned bilateral cataract extraction, and ability to adhere to study supplementation. Exclusion criteria included known retinal disease (e.g., diabetic retinopathy or retinal vein occlusion), prior intraocular surgery, or recent use of supplements containing TCA-cycle intermediates. Baseline AH was collected during the first-eye surgery. Participants then initiated oral Ca-AKG supplementation (2 g/day, once daily) beginning 7 days before their second-eye surgery and continued until the day of the procedure. A second AH sample (∼50 μL) was collected at the start of the contralateral surgery under identical sterile conditions and processed immediately for storage at -80 °C.**(Figure 2)**

The Ca-AKG dosage was selected based on prior safety data and preclinical efficacy studies. In murine models, dietary Ca-AKG at scaled-equivalent doses extended lifespan, reduced frailty, and improved mitochondrial and redox homeostasis without evidence of toxicity.^21,22^ Human nutraceutical studies have similarly demonstrated that oral Ca-AKG at 1-6 g/day is well tolerated, produces measurable increases in circulating α-KG, and engages metabolic pathways relevant to mitochondrial function.^23,24^ A 2 g/day regimen therefore represents a biologically active yet conservative dose expected to achieve systemic exposure sufficient to test whether α-KG traverses to the aqueous humor and modulates intraocular TCA-cycle metabolites in a short-term intervention.

### AH Metabolite Extraction

A total of 20 eye fluid samples were analyzed in this study, consisting of 10 samples collected at baseline and 10 paired samples collected after Ca-AKG supplementation. Metabolite extraction was conducted using methanol precipitation, following the standardized protocol provided by the Calgary Metabolomics Research Facility (CMRF). Samples were treated with 100% methanol and kept on ice for 30 minutes, with vortexing every 10 minutes. Centrifugation was performed at 21,000 × g for 10 minutes at 4 °C, after which the supernatant was transferred into clean microcentrifuge tubes and stored at −80 °C. Prior to submission, extracts were gently vortexed and centrifuged again at 21,000 × g for 10 minutes. From each sample, 20 µL of the supernatant was aliquoted into a 96-well microplate and diluted with 180 µL of 50% methanol before transfer to CMRF.

### Metabolomics Mass Spectrometry

Targeted quantitative metabolite analyses were performed using previously described protocols.^25–27^ Samples were analyzed on a Q Exactive™ HF Hybrid Quadrupole-Orbitrap™ Mass Spectrometer (Thermo Fisher), coupled to a Vanquish™ UHPLC System (Thermo Fisher). Separation of metabolites was achieved by ultra-high performance liquid chromatography (UHPLC) using a binary solvent system on a Syncronis HILIC UHPLC column (2.1 mm × 100 mm × 1.7 µm) at a flow rate of 600 µL/min. Solvent A was 20 nM ammonium formate in MS-grade water (pH 3.0), while solvent B consisted of MS-grade acetonitrile containing 0.1% formic acid (v/v). The gradient profile was: 0–2 min, 100% B; 2–7 min, 100–80% B; 7–10 min, 80–5% B; 10–12 min, 5% B; 12– 13 min, 5–100% B; 13–15 min, 100% B. A 2 µL injection volume was used, and acquisition was performed in negative ion mode with full scan, at a resolution of 240,000 across an m/z range of 50–750. Metabolite identification was based on comparison of experimental m/z values (±10 ppm) and retention times with authentic reference standards (MSLSTM Sigma-Aldrich). Peak annotation and data processing were performed with the El-MAVEN software package^28–30^. For quantification, metabolite levels were normalized against an eight-point calibration curve constructed from commercial standards. For quality control (QC), metabolites were required to meet strict peak-identification thresholds, including signal intensity >10□, mass-to-charge deviation <5 ppm, retention-time deviation <0.5 min, and a sample-to-blank ratio >10:1. Concentrations were further confirmed by standard-curve performance (R² ≥ 0.98) and linear-range compliance. Metabolites not meeting these thresholds, most commonly due to low signal intensity or insufficient sample-to-blank separation in AH, were excluded to ensure high-confidence quantification.

### Bioinformatics and Statistical Analysis

Normalized proteomics data were provided by Somalogic and imported to R Studio (version 2025.09.1+401, R version 4.5.1). Aptamers’ target annotation and mapping to UniProt accession numbers as well as Entrez gene identifiers were provided by Somalogic. The estimated limit of detection (eLoD) was calculated for each aptamer using a ‘robust estimate’ method as previously described.^31^ Gene ontology analysis using ShinyGO 0.85 was used to identify 695 mitochondrial proteins (GO:0005739), which underwent analysis using 1-way ANOVA ^32^. Differentially expressed mitochondrial proteins were determined using the limma package with default parameters except using method = “robust” in lmFit. P-values were corrected for multiple testing using Benjamini-Hochberg adjustment implemented in the limma package ^33^. Proteins with log2 fold change (log2FC) >0.5 or <-0.5 and p-value <0.05 were considered as differentially expressed proteins (DEP). Functional annotation of differentially expressed proteins was performed with Metascape (https://metascape.org/) ^34^. The STRING (Search Tool for the Retrieval of Interacting Genes) v11 database (https://string-db.org) was used to identify protein-protein interactions and illustrate interconnectivity among proteins ^35^. Data were analyzed at a false discovery rate (FDR) of 5% and the protein-protein interaction networks were analyzed.

Metabolomic analyses were performed in Python using the pandas, NumPy, SciPy, statsmodels, and matplotlib libraries. Pre- and post-supplementation metabolite values were treated as paired measurements and evaluated using two-tailed paired t-tests under the null hypothesis of no mean difference.

### Quantification and Statistical Analysis

Statistical parameters were reported either in individual figures or corresponding figure legends. Quantified data were in general presented as bar or line plots, with the error bar corresponding to mean ± SD. All statistical analyses were performed in R or GraphPad Prism. Whenever asterisks are used to indicate statistical significance, * stands for P < 0.05; ** for P < 0.01, and *** for P < 0.001.

## Results

### Dysregulation of mitochondrial and TCA-cycle proteins in GA

To characterize mitochondrial proteomic changes in GA, 20 patients diagnosed with GA were included in the analysis (mean age: 73.4 ± 5.1 years). For comparison, 10 individuals without retinal disease served as controls (mean age: 83.4 ± 9.2 years) (**Table 1**). Controls were older cataract surgery patients without clinical or imaging evidence of AMD; despite the lack of age matching, their advanced age provides a conservative reference for GA-associated alterations. AH samples were safely obtained from this pilot study cohort without complication and underwent targeted proteomic analysis to identify mitochondrial pathways as previously described^31^. A total of 64 mitochondrial proteins were differentially expressed among control and GA AH (p<0.05; **Fig. 1A**). Among the significantly downregulated mitochondrial proteins in GA were the TCA cycle-related proteins dihydrolipoyllysine-residue succinyltransferase (DLST), ornithine aminotransferase (OAT), pyruvate dehydrogenase E1 component subunit beta (PDHB), and succinate dehydrogenase [ubiquinone] iron-sulfur subunit (SDHB; **Fig. 1A**). Among the significantly upregulated mitochondrial proteins in GA were NADP-dependent malic enzyme (ME1), sorbitol dehydrogenase (SORD), 2-iminobutanoate/2-iminopropanoate deaminase (RIDA), glutathione reductase (GSR), and CDGSH iron-sulfur domain-containing protein 1 (CISD1; **Fig. 1A**).

**Figure 1.**
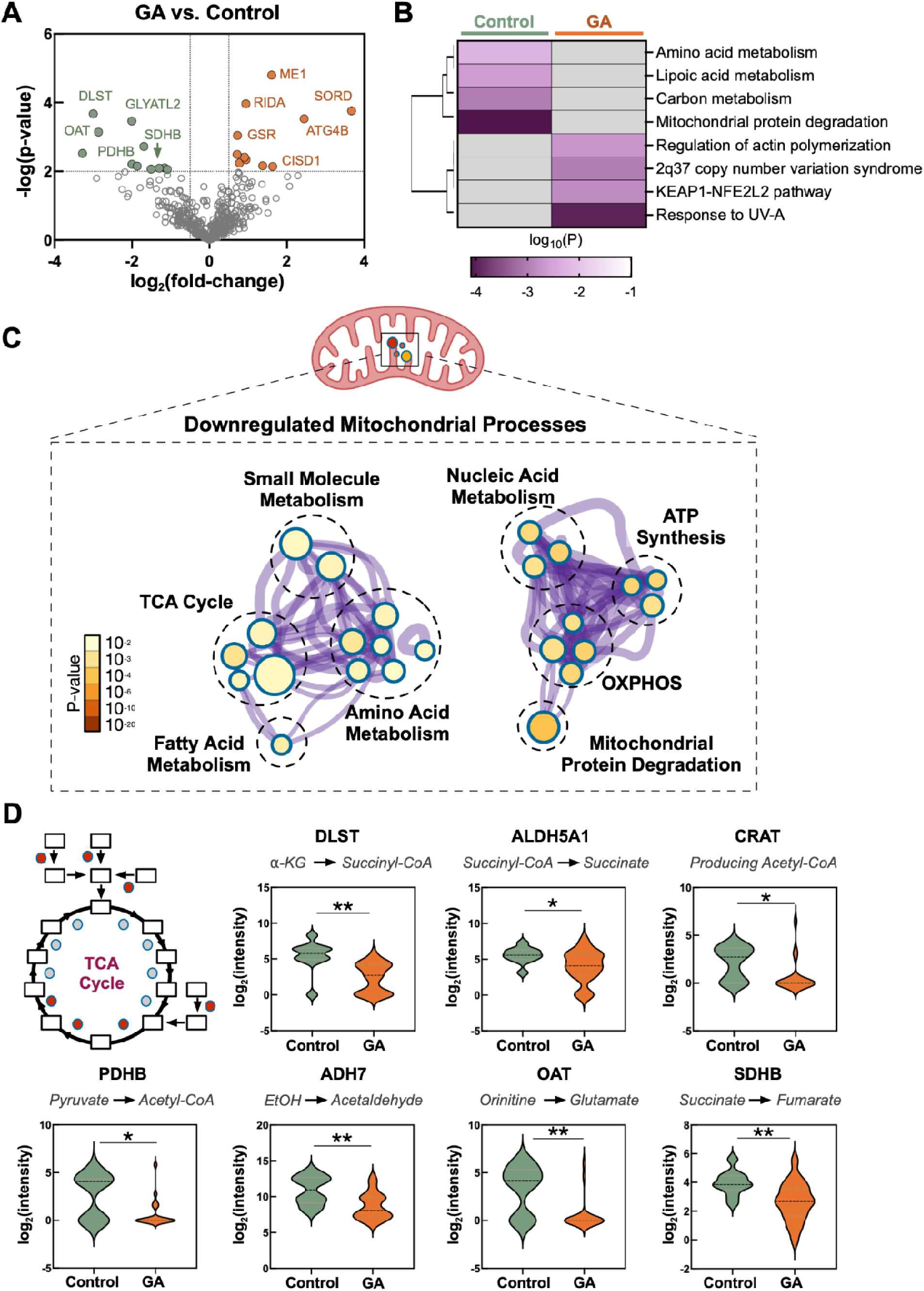
Proteomic signatures of GA reveal downregulation of TCA-cycle and mitochondrial pathways. **(A)** Differential protein abundance between GA and control aqueous humor samples was assessed using one-way ANOVA and visualized as a volcano plot. The x-axis shows log₂ fold change (GA vs. control), and the y-axis shows statistical significance expressed as −log₁₀(p-value). Selected mitochondrial and metabolic proteins are highlighted. **(B)** Pathway overrepresentation analysis of GA AH proteomics was performed using Metascape. Enriched pathways were identified using a two-sided hypergeometric test, filtered by accumulative p-values and enrichment factors, and hierarchically clustered based on Kappa-statistical similarity (threshold = 0.3). The upper cluster highlights pathways downregulated in GA relative to controls, including mitochondrial protein degradation, carbon metabolism, lipoic acid metabolism, and amino acid metabolism, while color intensity reflects pathway-level significance (log₁₀ p-value). **(C)** Network representation of significantly downregulated mitochondrial gene ontology terms in GA. Nodes represent enriched biological processes, colored by statistical significance, with larger clusters reflecting pathways containing a greater number of affected proteins. Major functional groupings include TCA cycle, oxidative phosphorylation (OXPHOS), amino acid metabolism, fatty acid metabolism, ATP synthesis, and mitochondrial protein degradation. **(D)** Schematic overview of the TCA cycle highlighting identified downregulated proteins (red circles; top left), alongside violin plots showing relative protein abundance of representative TCA- and mitochondrial-related enzymes in control versus GA samples. Statistical significance is indicated as *P < 0.05, **P < 0.01, and ***P < 0.001.

**Figure 2.**
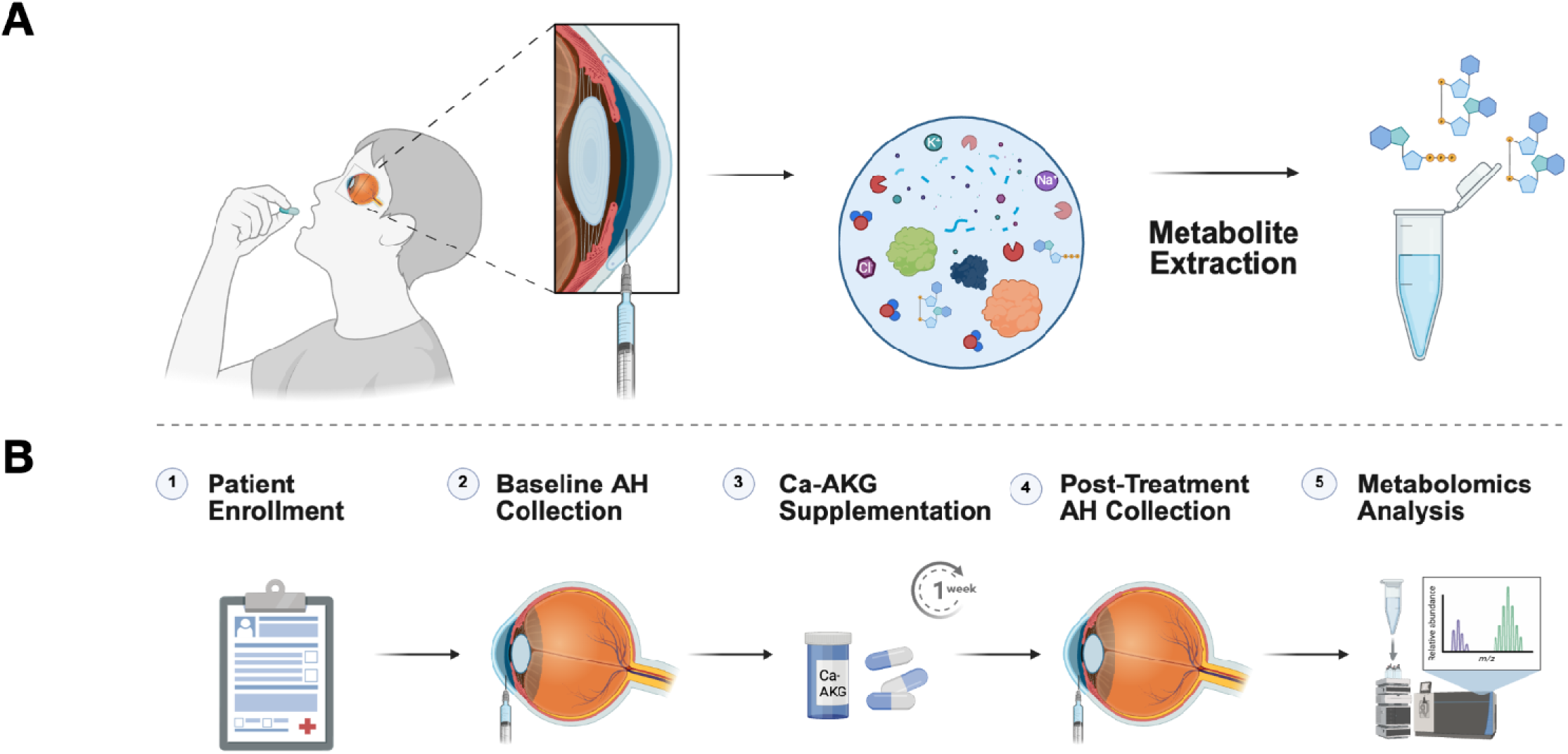
Study workflow for AH sampling, α-KG supplementation, and metabolomic analysis. **(A)** AH was collected from the anterior chamber during cataract surgery, followed by metabolite extraction and liquid chromatography–mass spectrometry (LC–MS) analysis. **(B)** Schematic overview of the Phase 0 interventional study design. Participants undergoing sequential bilateral cataract surgery provided paired AH samples, with baseline collection performed during first-eye surgery. Subjects then received oral Ca-AKG (2 g/day) for 7 days prior to second-eye surgery, at which time a post-intervention AH sample was collected. All specimens were analyzed using targeted metabolomics.

**Table 1.** Demographic characteristics for GA and control cohorts included in the proteomics discovery analysis.

|  | Geographic Atrophy Cohort (n = 20) | Control Cataract Cohort (n = 10) |
| --- | --- | --- |
| Age (mean ± SD) | 73.4 ± 5.1 | 83.4 ± 9.2 |
| Sex, n (F/M) | 8/12 | 8/2 |

To further classify these mitochondrial proteins in GA aqueous, we performed pathway analysis, which identifies groups of functionally-linked proteins. An enrichment of proteins associated with mitochondrial protein degradation (logP = -4.09), carbon metabolism (logP = -3.03), lipoic acid metabolism (logP = -2.65), and amino acid metabolism (logP = -2.29) was identified in control AH compared to GA (**Fig. 1B**). As an orthogonal validation analysis, using Metascape, we found many enriched biological process terms and 26 out of 30 downregulated mitochondrial proteins were included in a biological network (**supplemental figure 1**). Among the retrieved biological process terms, those related to *mitochondrial protein degradation* (9 proteins) and *carboxylic acid metabolic process* (8 proteins) were among the most significant. A subnetwork of mitochondrial and metabolic pathway terms represented among the downregulated proteins is depicted in **Fig. 1C**. Taken together, these results indicated that mitochondrial metabolic pathways, particularly the proteins implicated in the TCA cycle, were disrupted in GA-affected eyes (**Fig. 1D**).

### α-KG Uptake in the Eye and Coordinated Metabolic Shifts

Building on the proteomic evidence of TCA-cycle protein depletion in GA, we next asked in an open-label, single-arm, phase 0 study (i) whether systemic supplementation could directly influence intraocular TCA-cycle activity in living human eyes, and (ii) whether such changes could be detected in AH. A separate cohort of 8 patients scheduled for bilateral sequential cataract surgery was enrolled, allowing AH collection from both first- and second-eye procedures in these patients and yielding 16 paired samples.The cohort had a mean age of 70.1 years (range 56–79), was evenly distributed by sex (4 females and 4 males), and had a mean BMI of 26.3 (range 21–37) **(Table 2)**. All predefined adverse events (AE) were prospectively monitored across gastrointestinal, metabolic/electrolyte, renal/urinary, neurologic, ophthalmic, cardiovascular, and systemic categories, as well as serious AE. No AE were observed in any category, and no patient experienced symptoms such as gastrointestinal discomfort, electrolyte disturbances, renal changes, neurologic complaints, ophthalmic inflammation or IOP elevation, cardiovascular changes, allergic reactions, or any life-threatening events during Ca-AKG supplementation **(Table 3)**.

**Table 2.** Baseline demographic and clinical characteristics of participants enrolled in the Ca-AKG supplementation study.

| Patient | Sex | BMI | Medical History | Ocular History |
| --- | --- | --- | --- | --- |
| 1 | F | 21 | Osteoporosis | Dry eye disease |
| 2 | F | 23 | Osteoporosis | N/A |
| 3 | M | 29 | Hypertension, Hyperlipidemia | N/A |
| 4 | M | 22 | Hyperlipidemia, Benign prostatic hyperplasia | N/A |
| 5 | M | 29 | Obstructive sleep apnea | Early AMD |
| 6 | M | 27 | Osteoporosis | N/A |
| 7 | F | 37 | Osteoporosis, Obesity, Prediabetes, Chronic obstructive pulmonary disease | Epiretinal membrane |
| 8 | F | 22 | Osteoporosis | Early AMD |

\* BMI, body mass index (kg/m<sup>2</sup>); AMD, age-related macular degeneration; N/A indicates no reported ocular disease; All participants underwent sequential bilateral cataract surgery.

**Table 3.** Treatment-emergent adverse events following oral Ca-AKG supplementation in the Phase 0 cohort.

| Adverse Event Category | Specific Events Monitored | Number of Events (n) |
| --- | --- | --- |
| Gastrointestinal | Nausea, abdominal discomfort, bloating, diarrhea, constipation | 0 |
| Metabolic / Electrolyte | Hypercalcemia, hypocalcemia, metabolic alkalosis, electrolyte abnormalities | 0 |
| Renal / Urinary | Changes in serum creatinine or eGFR, kidney discomfort, nephrolithiasis symptoms | 0 |
| Neurologic | Headache, dizziness, fatigue | 0 |
| Ophthalmic | IOP elevation, inflammation, ocular discomfort | 0 |
| Cardiovascular | Blood pressure changes, palpitations | 0 |
| General / Systemic | Rash, allergic reaction, malaise | 0 |
| Serious Adverse Events (SAE) | Hospitalization, life-threatening event | 0 |

\* Values represent the total number of treatment-emergent adverse events observed during the supplementation period and perioperative follow-up. No adverse events were reported by any participant. eGFR, estimated glomerular filtration rate; IOP, intraocular pressure.

We performed a targeted metabolomics analysis of 100 metabolites, of which 47 met QC criteria and were retained for downstream analysis **(Table S1)**. Among these, several TCA-cycle intermediates, including α-KG and succinate, as well as related metabolites such as glutamine and acetoacetate were quantified, enabling pathway-level assessment of metabolic changes following supplementation **(Fig 3A)**. In the metabolic analysis, seven participants exhibited concordant, significant increases in α-KG following supplementation (p.adj=0.0091). These findings indicate that orally administered α-KG is detectable in the AH and is associated with increased α-KG levels in most participants. Only one participant (Patient 7) demonstrated a decrease in α-KG and was noted to have significant systemic metabolic comorbidities, including prediabetes and chronic obstructive pulmonary disease, as well as a BMI of 37, which meets WHO and CDC criteria for obesity (BMI≥30 kg/m²) and metabolic syndrome. Given these factors, this participant was excluded from subsequent analysis. **(Fig 3B)**

**Figure 3.**
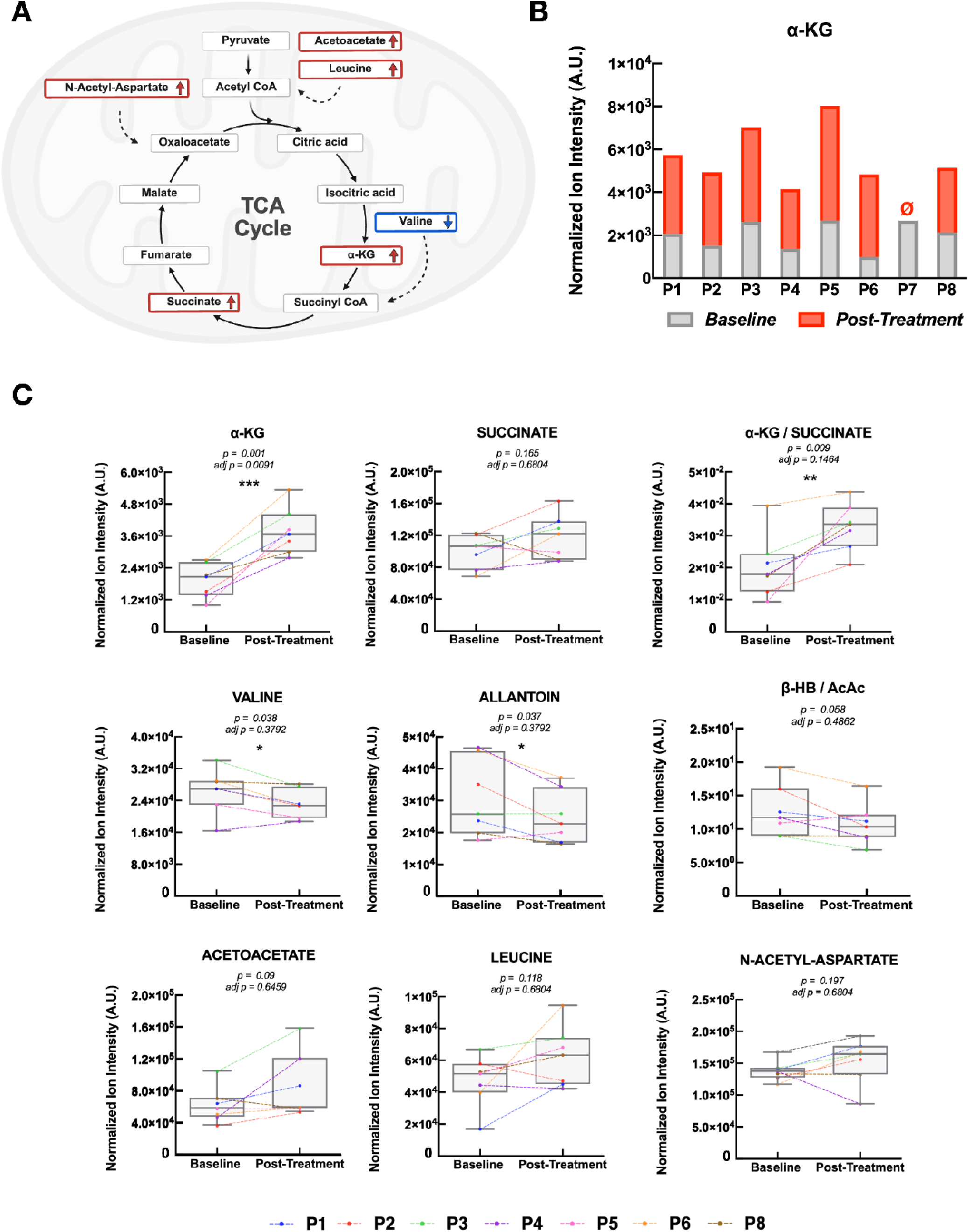
Intraocular TCA-cycle–related metabolite responses to oral α-KG supplementation. **(A)** Schematic overview of the TCA cycle highlighting metabolites quantified in AH. Metabolites demonstrating a post-treatment increase are indicated in red, while metabolites showing a decrease are indicated in blue, based on paired pre-/post-supplementation comparisons. **(B)** Individual AH α-KG levels measured at baseline (grey) and after oral Ca-AKG supplementation (red). Each bar represents normalized ion intensity for an individual participant, demonstrating a consistent post-treatment increase in intraocular α-KG across subjects, with one metabolic non-responder indicated (Ø). **(C)** Paired pre- and post-treatment metabolite trajectories for TCA-cycle intermediates and related metabolites following 7 days of Ca-AKG supplementation. Boxplots show group distributions with overlaid colored lines representing individual participant trajectories. Metabolites shown include α-KG, succinate, α-KG/succinate, valine, acetoacetate, leucine, N-acetyl-aspartate, allantoin, and the β-hydroxybutyrate/acetoacetate (β-HB/AcAc) ratio, a surrogate marker of mitochondrial NADH/NAD redox state. Statistical significance was assessed using paired t-tests, with nominal and multiple-testing-adjusted p-values indicated for each metabolite (*P < 0.05, **P < 0.01, ***P < 0.001). Collectively, these data demonstrate coordinated modulation of TCA-cycle-related metabolites and redox-associated markers following systemic α-KG supplementation.

In the remaining seven patients, the α-KG-to-succinate ratio (α-KG/succinate), an established indicator of TCA-cycle activity, was increased (p=0.009) following supplementation. Because this ratio reflects the balance between an upstream TCA-cycle intermediate and its downstream product, it provides a pathway-level metric that is less influenced by absolute metabolite concentrations and more reflective of relative TCA-cycle flux .^36^ The observed increase in α-KG/succinate elevation likely indicates enhanced TCA-cycle activity rather than simple accumulation of the supplemented precursor.

Another important metric, the β-hydroxybutyrate-to-acetoacetate ratio (β-HB/AcAc), a commonly used indicator of the mitochondrial redox state, showed a trend toward reduction following supplementation (p=0.058). Because higher β-HB/AcAc ratios reflect a more reduced mitochondrial state, this observation may be consistent with a shift toward enhanced mitochondrial redox balance rather than simple ketone accumulation. In parallel, allantoin, a validated marker of oxidative stress in humans^37^, was reduced (p=0.037), suggesting a potential decrease in oxidative burden following supplementation. Consistent with these broader shifts in mitochondrial energy metabolism, valine, a branched chain amino acid that feeds into the TCA cycle, was also reduced (p=0.038), suggesting altered amino acid utilization in the context of t mitochondrial metabolism following α-KG supplementation **(Fig 3C)**. While unadjusted analyses identified nominally significant differences (p < 0.05), these effects did not remain significant after multiple-comparison adjustment and should be interpreted as exploratory, pending confirmation in larger, adequately powered studies.

Among the other metabolites analyzed, excluding α-KG and closely related energy-associated metabolites, no widespread changes were detected in amino acid, nucleotide, carbohydrate, or vitamin-related pathways following Ca-AKG supplementation. This suggests that oral Ca-AKG induces a highly targeted metabolic effect without broadly perturbing unrelated metabolic pathways.

## Discussion

We demonstrate that oral α-KG supplementation induces quantifiable and biologically coherent changes in the intraocular metabolic profile of living humans. In the first cohort, we observed reduced TCA-cycle proteins in GA patients, consistent with impaired mitochondrial metabolic capacity. In a second, interventional cohort, we showed that oral α-KG can be safely administered and enhances TCA-cycle metabolism in the human eye. Together, these findings provide direct in vivo evidence that the intraocular metabolic state is dynamically responsive to systemic metabolic inputs and reveal metabolic plasticity in the living human eye.

From a translational standpoint, this study further establishes the procedural and bioinformatic feasibility of deep multi-omics profiling from minimal AH volumes obtained during routine ophthalmic surgery procedures. Although AH sampling is intrinsically volume-limited, recent advances in mass spectrometry sensitivity and pathway-level bioinformatic integration now allow meaningful proteomic and metabolomic inference from ten to fifty microliters of ocular fluid. Importantly, these modalities provide complementary, non-redundant information: proteomics reflects longer-term alterations in pathway capacity, while metabolomics captures dynamic, intervention-responsive flux. Their integration enables a more comprehensive and clinically actionable view of intraocular metabolism than either modality alone. Our study establishes the feasibility of multi-omics liquid biopsies to capture comprehensive molecular data on enzyme behavior in living human eyes, something previously only possible in tissue culture or animal models.

Importantly, our design also aligns with a phase 0 (exploratory) clinical-trial strategy, which is particularly valuable in ocular metabolism where efficacy trials are slow, costly, and often confounded by heterogeneous disease biology. Phase 0 studies are explicitly intended to de-risk translation early by demonstrating (i) human bioavailability at the relevant site, (ii) target engagement / pharmacodynamic modulation, and (iii) assay feasibility using small cohorts and limited exposure, enabling faster decisions before launching outcome-powered trials.^38^ In the context of GA, this is critical because the central uncertainty is not only whether α-KG has systemic effects, but whether a systemic metabolic input can penetrate the eye and measurably shift intraocular mitochondrial pathways in vivo, a question that cannot be answered reliably from post-mortem tissue or animal models alone.

Over the years, α-KG has attracted growing scientific attention and evolved from a simple TCA cycle intermediate into a key integrator of mitochondrial function. In a seminal study, Chin et al. showed that dietary α-KG supplementation extends lifespan and delays frailty in mice by inhibiting ATP synthase and suppressing mTOR signaling.^18^ Asadi Shahmirzadi et al. further demonstrated that Ca-AKG supplementation in middle-aged mice not only prolongs lifespan but also compresses morbidity and attenuates chronic inflammation via induction of the anti-inflammatory cytokine IL-10.^21^ Consistent with these systemic effects, our group previously reported that oral α-KG supplementation preserves visual function and protects photoreceptors in a mouse model of retinitis pigmentosa, supporting a role for α-KG in sustaining mitochondrial metabolism and neuronal survival within the retina.^17^ These studies position α-KG as a key regulator in linking mitochondrial metabolism, inflammatory signaling, and neuroprotection across aging and retinal disease. Beyond preclinical work, α-KG has begun to be explored in human aging studies and clinical trials. In a recent trial of 42 healthy adults, sustained-release α-KG administration for approximately seven months led to a mean reduction of nearly eight years in biological age, as measured by DNA methylation clocks.^39^ While epigenetic aging clocks are not readily applicable to ocular tissues, we have developed a liquid-biopsy-based proteomic clock for the human eye.^31^ This platform enables direct assessment of ocular biological age using AH proteomic data obtained on the same analytical platform used to quantify mitochondrial proteins. Such an approach creates a unique opportunity to test the hypothesis that mitochondrial-targeted metabolic interventions, including α-KG supplementation, can modulate the biological age of the human eye in vivo.

α-KG’s central role in regulating cellular metabolism is particularly important in the eye, where mitochondrial dysfunction has emerged as a shared pathogenic mechanism across multiple ocular diseases and reflects the eye’s extraordinary bioenergetic demands.^40^ Photoreceptors, retinal pigment epithelial (RPE) cells, Müller glia, and retinal ganglion cell axons depend on continuous oxidative phosphorylation to sustain ion transport, synaptic signaling, and photoreceptor outer-segment renewal.^41^ Mitochondria coordinate substrate oxidation and redox balance to meet these demands, and even subtle impairments can destabilize retinal homeostasis, leading to oxidative stress, inflammation, and neurodegeneration.^42,43^ In AMD postmortem tissue, Feher et al. demonstrated reduced RPE mitochondrial content and disrupted cristae structure, while Decanini et al. and Nordgaard et al. similarly reported oxidative damage and dysregulated mitochondrial biogenesis with disease progression.^44–46^ In diabetic retinopathy, persistent oxidative stress disrupts mitochondrial function, and Kowluru et al. showed that resulting cytochrome c release and Bax translocation drive retinal capillary cell apoptosis^47^, and Madsen-Bouterse et al. observed mitochondrial DNA (mtDNA) damage persisting despite glycemic control being restored.^48^ In glaucoma, Abu Amero et al. found ∼21% lower mitochondrial respiratory activity and novel mtDNA variants compared to age-matched controls.^49^ Together, these findings reveal a shared metabolic vulnerability in ocular tissues reliant on uninterrupted oxidative metabolism, positioning mitochondrial insufficiency as a central and potentially tractable driver of vision loss. However, these observations have been derived primarily from post-mortem human tissues, and none have directly examined GA in living human eyes using liquid biopsy approaches. Previous lines of evidence suggest that systemic α- KG could theoretically reach the eye, including the presence of high-affinity monocarboxylate transporters on the RPE^50,51^, the ability of α-KG to cross the blood-brain barrier, and the central role of reductive carboxylation in RPE metabolism. Our multi-omics liquid biopsy platform can now confirm this in individual human eyes.

Using AH liquid biopsies from living patients with GA, we identified a pronounced reduction in mitochondrial proteins and a coordinated suppression of oxidative metabolic pathways. Pathway and ontology analyses further identified mitochondrial protein degradation, carboxylic acid metabolism, and oxidative bioenergetic processes as the most significantly affected pathways in GA. Together, these findings implicate mitochondrial insufficiency and disrupted TCA-cycle activity as central metabolic targets in retinal degeneration, providing the rationale to test whether oral supplementation could support restoration of these impaired pathways. This approach establishes feasibility, enables quantitative assessment of metabolic changes in the living human eye, and lays the groundwork for future interventional clinical trials in GA.

Within this context, we evaluated whether α-KG, a central intermediate of the TCA cycle, could reach the eye and modulate intraocular metabolism. We observed a significant elevation of α-KG in AH following supplementation, indicating systemic bioavailability and effective penetration into the eye. Beyond sustaining TCA-cycle flux and anaplerosis, α-KG serves as a metabolic signal linking mitochondrial energy production with epigenetic and hypoxia-responsive regulation.^52,53^ Its increase in AH therefore supports the capacity of systemic metabolic interventions to directly influence intraocular mitochondrial pathways.

The coordinated increase in α-KG together with an increased α-KG/succinate ratio, rather than an isolated rise in succinate, supports enhanced forward TCA-cycle flux with preserved oxidative phosphorylation, rather than passive accumulation of supplemented substrate. Succinate occupies a unique metabolic position as the only measured metabolite that directly couples the TCA cycle to the electron transport chain (ETC). When mitochondrial respiration is impaired, succinate tends to accumulate, lowering the α-KG/succinate ratio; in contrast, the pattern observed here is consistent with efficient succinate oxidation and intact TCA-ETC coupling. Given prior evidence that retinal succinate fuels RPE mitochondrial respiration and oxygen consumption,^54,55^ these findings provide a mechanistically direct link between α-KG supplementation and improved oxidative metabolism in the outer retina, reinforcing mitochondrial dysfunction as a tractable target in GA.

In line with these flux-based findings, pathway-level metabolic ratios provided additional insight into mitochondrial redox balance following α-KG supplementation. The β-hydroxybutyrate/acetoacetate (β-HB/AcAc) ratio, a commonly used indicator of mitochondrial NADH/NAD⁺ redox state, showed a consistent downward trend, indicating more efficient electron transfer and improved oxidative metabolism.^56^ Concurrently, allantoin, a well-validated marker of oxidative stress in human biofluid, was also significantly reduced, suggesting a systemic reduction in oxidative burden following α-KG supplementation.^37,57^ Together, these concordant changes support a coordinated improvement in mitochondrial metabolic balance rather than isolated metabolite effects. Importantly, when viewed alongside the mitochondrial pathway suppression we observed in GA patients, these α-KG responsive redox and improved stress markers indicate that ocular mitochondrial metabolism remains dynamically responsive to systemic metabolic supplementation, supporting the biological feasibility of future α-KG-based interventional trials in GA.

In this study, we used sequential AH samples obtained during first- and second-eye cataract surgery to study metabolic changes after oral α-KG supplementation. This approach is supported by evidence that baseline inter-eye variability in AH metabolomics is negligible. In a simultaneous bilateral cataract surgery study, Pietrowska et al. demonstrated that AH metabolomic profiles show high interocular similarity, with a single eye broadly representative of the fellow eye for most metabolites, with only taurine differing significantly.^58^ In contrast, Li et al. reported metabolic differences in the second eye during sequential surgery, but their inter-surgery interval was less than two weeks, and a subsequent commentary noted that these changes may reflect early postoperative or surgery-related effects rather than true biological asymmetry.^59,60^ Given the four-week interval in our study and the established low intrinsic inter-eye variability, differences observed in our cohort are more likely to represent true biological changes, supporting the validity of our paired-eye analytical framework. Nonetheless, future studies incorporating pre- and post-intervention AH sampling would further strengthen causal inference, particularly in diseases like AMD where disease burden and stage can vary significantly between eyes.

The identification of a metabolic non-responder (Patient 7) further highlights the practical value of liquid biopsy in interventional clinical trials. Unlike genetic markers, which remain fixed, metabolic profiles reflect dynamic systemic states (e.g. obesity, insulin resistance) that may influence ocular drug delivery and metabolic responsiveness. These observations suggest that metabolic therapies for AMD may benefit from localized “metabolic phenotyping” to inform patient selection and dosing, rather than relying on a one-size-fits-all approach. Systemic metabolic comorbidities may therefore represent an important source of biological heterogeneity and should be considered in the design or exclusion criteria of future clinical trials.

### Study Limitations

Despite these exciting preliminary mechanistic findings, this study has some limitations. First, the discovery cohort was relatively small and the supplementation interval shorter than typical systemic aging-intervention regimens. However, the paired-eye design enhanced analytical sensitivity by allowing each participant to serve as their own internal control, and significant metabolic changes were still detectable within this limited timeframe. In addition, our targeted metabolomics panel assessed only a subset of metabolites, though it was sufficient to detect consistent TCA-related shifts after α-KG supplementation; future studies with broader untargeted analyses may help contextualize these changes within the global metabolic network. While our collection protocol for proteomics is well established, the metabolic collection profile could be improved by immediate extraction and shorter intervals before analysis that we expect will likely stabilize and improve metabolite measurement. Finally, although we observed consistent changes in TCA-cycle metabolites, how these biochemical shifts translate into functional or phenotypic improvements remains uncharacterized and will require follow-up longitudinal studies and interventional trials that incorporate clinical outcomes. Multiple α-KG formulations are currently available and have been evaluated in human studies, including sustained-release Ca-AKG combined with micronutrients.^39^ These formulations will enable future studies to determine the optimal dosing, formulation, and exposure duration that yield the greatest biological and clinical effects. Taken together, despite these limitations, this study still provides a robust proof-of-concept that systemic metabolic interventions can dynamically modulate intraocular metabolism in living humans.

## Conclusions

In summary, this study demonstrates that liquid-biopsy proteomics can detect disrupted metabolic pathways in living human eyes with GA, and that the intraocular metabolism can be directly perturbed and measured *in vivo* using liquid-biopsy metabolomics. Oral α-KG supplementation induced coordinated changes in TCA-cycle metabolites, indicating that mitochondrial activity in the eye remains closely linked to whole-body metabolic state. When viewed alongside the mitochondrial pathway suppression in GA, these results reveal preserved metabolic plasticity in the human eye and provide a mechanistic foundation for targeting ocular energy metabolism through systemic interventions.

## Article Information

## Data Availability

All data produced in the present study are available upon reasonable request to the authors

## Acknowledgements

Metabolomics data were acquired at the Calgary Metabolomics Research Facility (CMRF), which is supported by the Alberta Center for Advanced Diagnostics.

## Author Contributions Statement

Dr. Mahajan had full access to all the data in the study and took responsibility for the integrity of the data and the accuracy of the data analysis. Study concept and design: TCY, GV, DKR, VBM. Acquisition of data: MZD, AK, AM, AD, VBM. Surgical sample collection: AM, SS, SS, AK, AMS. Analysis and interpretation of data: TCY, GV, SHL, AP, MC, AD, VBM. Drafting of the manuscript: TCY, VBM. Critical revision of the manuscript for important intellectual content: all authors. Statistical analysis: TCY, GV, AP. Obtained funding: VBM, PM. Administrative, technical, and material support: AGB, DA, PM, VBM. Study supervision: VBM.

## Institutional Review Board Statement

The study was approved by the Stanford University Institutional Review Board and adhered to the tenets set forth in the Declaration of Helsinki.

## Financial Support

TCY is supported by NIH grant 2T32EY027816. VBM is supported by NIH grants (R01EY031952, R01EY031360, R01EY030151, and P30EY026877), and Research to Prevent Blindness, New York, New York. GV is supported by NIH grant R38EY037090.

## Declarationh of Interests

Conflict of Interest Disclosures: None reported. Role of the Sponsor: The funding organizations had no role in design and conduct of the study; collection, management, analysis, and interpretation of the data; preparation, review, or approval of the manuscript; and decision to submit the manuscript for publication.

## Notes

### Competing Interest Statement

The authors have declared no competing interest.

### Clinical Trial

NCT07269704

### Author Declarations

All study procedures were approved by the Stanford University Institutional Review Board, complied with HIPAA regulations, adhered to the Declaration of Helsinki, and written informed consent was obtained from all participants.

